# The fragility index in randomised controlled trials of interventions for aneurysmal subarachnoid haemorrhage: a systematic review

**DOI:** 10.1101/2021.02.05.21251154

**Authors:** Aravind V Ramesh, Henry N P Munby, Matt Thomas

## Abstract

**Background:** Fragility analysis supplements the p-value and risk of bias assessment in the interpretation of results of randomised controlled trials. In this systematic review we determine the fragility index (FI) and fragility quotient (FQ) of randomized trials in aneurysmal subarachnoid haemorrhage.

**Methods:** This is a systematic review registered with PROSPERO (ID: CRD42020173604). Randomised controlled trials in adults with aneurysmal subarachnoid haemorrhage were analysed if they reported a statistically significant primary outcome of mortality, function (e.g. modified Rankin Scale), vasospasm or delayed neurological deterioration.

**Results:** We identified 3809 records with 17 randomized trials selected for analysis. The median fragility index was 3 (inter-quartile range 0-5) and the median fragility quotient was 0.012 (IQR 0-0.034). Six of nineteen trial outcomes (31.6%) had an fragility index of 0. In seven trials (36.8%), the number of participants lost to follow-up was greater than or equal to the fragility index. Only 17.6% of trials are at low risk of bias.

**Conclusions:** Randomised controlled trial evidence supporting management of aneurysmal subarachnoid haemorrhage is weaker than indicated by conventional analysis using p-values alone. Increased use of fragility analysis by clinicians and researchers could improve the translation of evidence to practice.

## Introduction

“The history of subarachnoid haemorrhage is similar to other areas of medicine in which anecdote leads to adoption of management that is of unproven e?cacy and safety until shown in high-quality randomised trials”[1]. The minimum standard of proof of efficacy and safety in traditional frequentist analysis is usually taken to be a p value of less than 0.05, subject to further interpretation of the entire body of evidence from the trial in question. However, if the p-value threshold for statistical significance is not met it is unlikely that the intervention studied will be adopted by clinicians[2,3].

What is less often appreciated is that crossing the arbitrarily defined threshold for statistical significance, and hence the minimum standard for adoption into clinical practice, may rest on as few as one or two actual events even in high quality randomised trials[4,5]. The number of events that need to change groups to render a statistically significant result non-significant is an indicator of the fragility of that result[6]. The “fragility index” is intended to supplement rather than replace the p-value; it may caution against the over simple classification of trials into positive and negative, and for positive trials, may offer an additional tool to gauge the strength of a result.

Previous application of fragility analysis to the field of neurocritical care has been limited. A review of cerebrovascular studies limited by date and database showed the median fragility index (FI) of seven randomised controlled trials (RCTs) in aneurysmal subarachnoid haemorrhage (aSAH) to be 5[7]. No studies of nimodipine, the only drug demonstrated to have a neuroprotective effect in SAH[1], were included in the review. Although the authors proposed a system of classification of RCTs using fragility index, no assessment of the risk of bias of the primary study, another key component of study appraisal, was included.

Our hypothesis is that RCT evidence supporting clinical practice in aneurysmal subarachnoid haemorrhage may be fragile. Our aim in this systematic review is to determine the Fragility Index of randomised controlled trials of interventions in aneurysmal subarachnoid haemorrhage that report a statistically significant patient centred primary outcome. The primary objective is to present FI for trials meeting the criteria described. Secondary objectives are to analyse the risk of bias, to compare the Fragility Index with the number of participants lost to follow up in the trial, and to calculate the Fragility Quotient (FQ)[8].

## Methods

This systematic review and meta-analysis was conducted following the ‘Preferred Reporting Items for a Systematic Review and Meta-analysis’ (PRISMA) statement[9] and the Synthesis Without Meta-analysis (SWiM) reporting guideline[10]. It was prospectively registered on the International Prospective Register of Systematic Reviews (PROSPERO 2020 – ID: CRD42020173604).

### Data sources and search strategy

The databases of Cochrane Central, MEDLINE, EMBASE, CINAHL and the trials registries of ClinicalTrials.gov and the World Health Organisation International Clinical Trials Registry Platform (WHO ICTRP) were searched on 11 March 2020 to identify studies meeting the eligibility criteria below. The search strategy (Supplemental Table 1) was designed by an academic librarian in MEDLINE without date or language restrictions, and then adapted to other databases.

### Eligibility Criteria

We included randomised clinical trials of acute hospital therapeutic interventions in adults older than 18 years of age with confirmed diagnosis of aneurysmal subarachnoid haemorrhage (aSAH) on neuroimaging or CSF analysis which met the following criteria: parallel group 1:1 randomisation, statistically significant dichotomous primary outcome (defined as p<0.05) that was one of mortality, functional outcome (e.g. dichotomised modified Rankin Scale or Glasgow Outcome Scale), vasospasm or delayed neurological deterioration. Where an outcome was reported at more than one time point the latest reported time point was used for analysis. Where available the intention to treat analysis of primary outcome was used to determine statistical significance for the purpose of inclusion.

Studies without an English language abstract reporting the primary outcome, or of traumatic subarachnoid haemorrhage or animals were excluded. Studies of diagnosis, rehabilitation or organisational aspects of aSAH management were excluded. Studies available only on pre-print servers rather than peer reviewed journals were excluded.

### Study selection

Two reviewers screened the titles and abstracts of articles. The full texts of potentially eligible studies were independently assessed by two reviewers; disagreements in each case were resolved through referral to the third reviewer.

### Data extraction and study appraisal

Two reviewers independently extracted data from eligible articles using a standardised data extraction form with discrepancies resolved by the third reviewer.

Risk of bias assessment for each study was performed independently by two reviewers with disagreements resolved by the third reviewer. The Cochrane Risk of Bias tool 2.0[11] was used to assess methodological quality.

### Data synthesis and analysis

The Fragility Index (FI) and Fragility Quotient (FQ) were calculated for each study as previously described[6,8].

The median (and interquartile range) value for FI is given for all studies meeting inclusion criteria and also by studies grouped according to intervention and primary outcome.

No quantitative data synthesis (meta-analysis) or further subgroup analysis was performed. All statistical analysis was performed using GraphPad Prism version 9.0.0 for Windows.

## Results

3809 citations were initially identified by the search strategy, with 17 studies involving 4151 participants included in qualitative data analysis (Figure 1).

**Figure 1:**
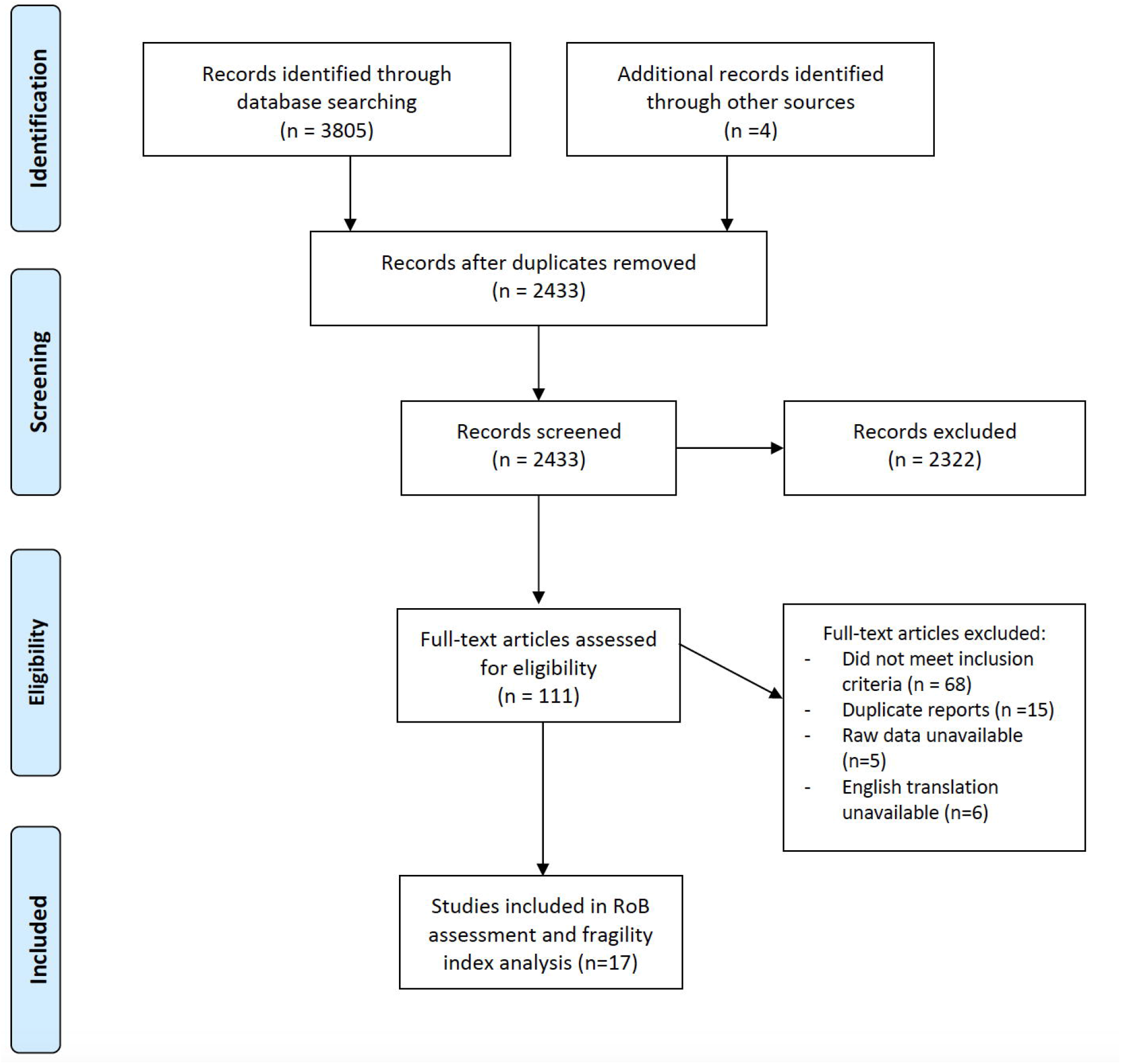
Preferred Reporting Items for Systematic Reviews and Meta-analyses flow diagram showing study selection for inclusion in Fragility analysis.

### Study characteristics

Study and participant characteristics are provided in Supplemental Table 2. Settings included Europe (six studies)[11–16], Asia (eight studies)[17–24], North America (two studies)[15,25], South America (one study)[26] and North Africa (one study)[27]. There were nine single centre studies[13,14,16,17,23–27] and eight multi-centre studies[11,12,15,18–22]. Publication dates ranged from 1989 to 2019. Thirteen studies looked at pharmacological interventions for aSAH[11,12,25–27,14,16–19,21–23]; the other four studies looked at surgical or interventional radiology treatments[15,20,24]. Eleven studies used vasospasm (angiographic or symptomatic) as their primary outcome[11,16,27,17–20,22,24–26], four studies used delayed ischaemic neurological deficit (DIND) or cerebral infarction[12–14,21] and three studies looked at clinical outcome using the Glasgow outcome scale (GOS) or the modified Rankin scale (mRS)[15,18,23].

### Fragility Index analysis

Nineteen outcomes across the seventeen studies met criteria for fragility index (FI) analysis (Table 1). The median FI for all trial primary outcomes was 3 (IQR 0-5). Six trial outcomes (31.6%) had an FI of 0. Only six trial outcomes (31.5%) had a FI of greater than three (Figure 2). The median fragility quotient (FQ) was 0.012 (IQR 0-0.034).

**Table 1:**
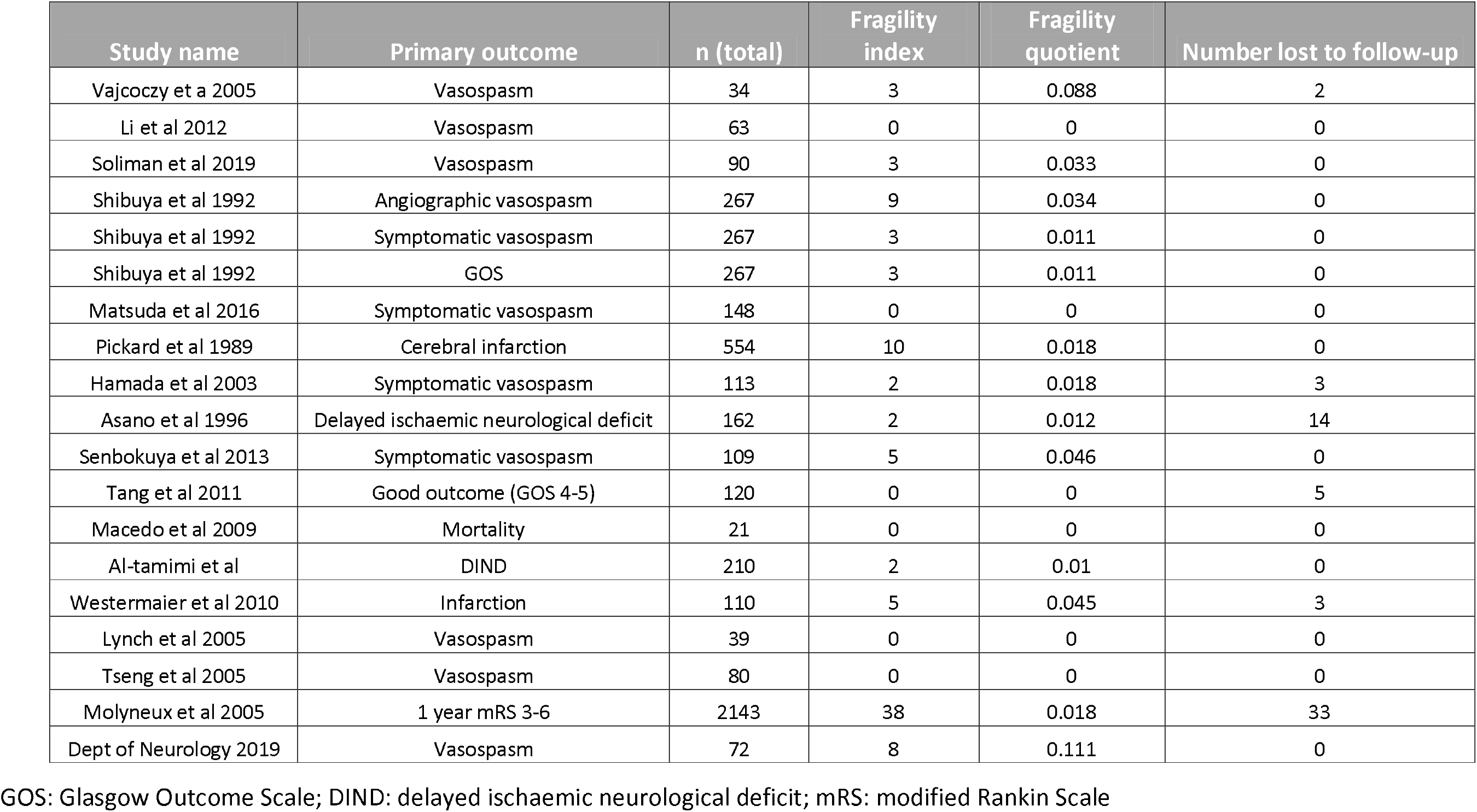
Fragility index, fragility quotient and number lost to follow up for included trials

**Figure 2:**
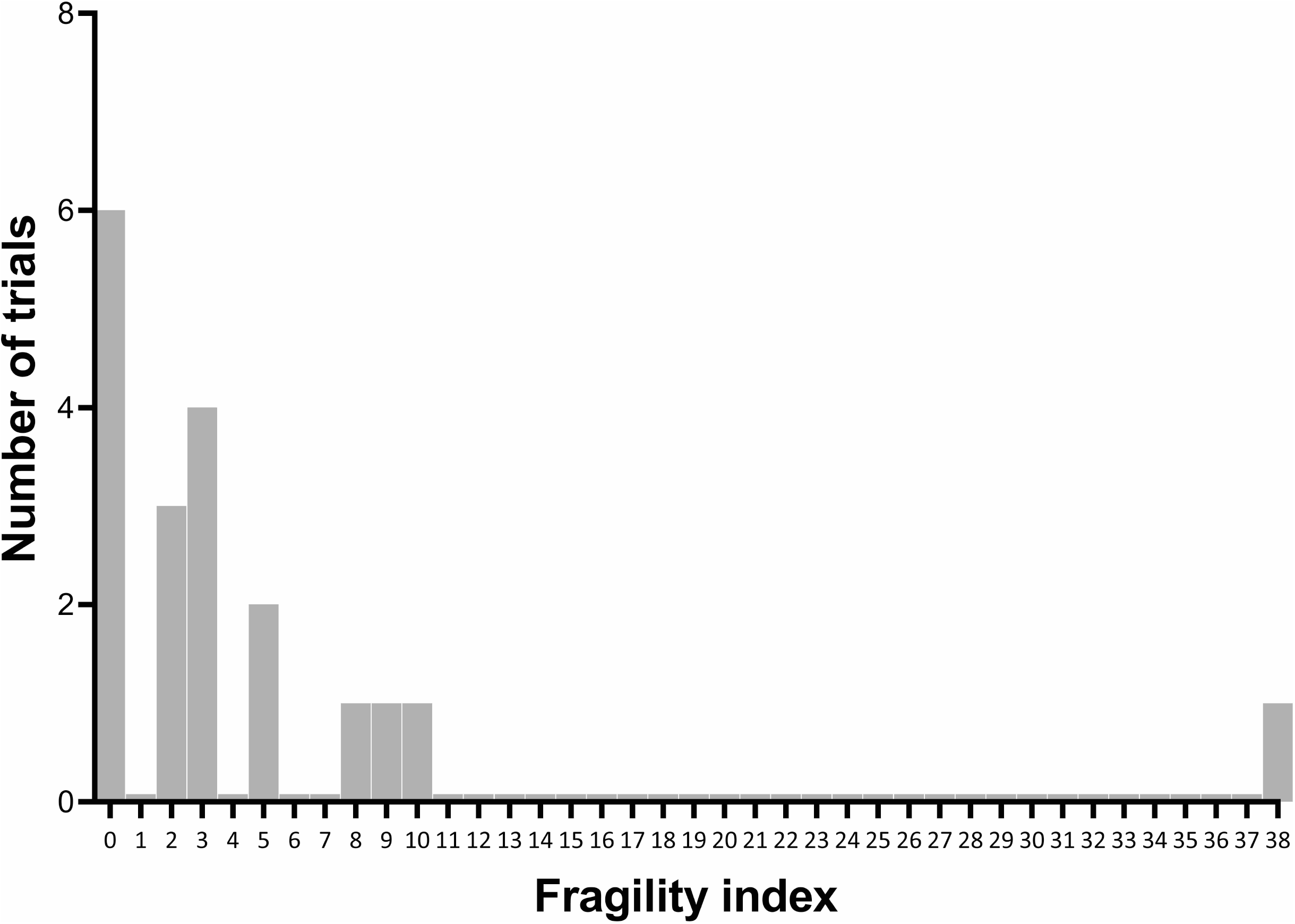
Distribution of fragility index of randomized controlled trials in aneurysmal subarachnoid haemorrhage reporting significant effects of an intervention on a prespecified patient-centred outcome.

Median trial size was 110 participants (IQR 67.5-186). In seven trials (36.8%), the number of participants lost to follow-up was greater than or equal to the fragility index.

The median FI for Vasospasm (both angiographic and symptomatic) was also 3 (11 trials, IQR 0-5), with a median FQ of 0.018 (0-0.046). The only trial reporting mortality as a primary outcome had an FI of 0. Four trials looked at delayed ischaemic neurological deficit (DIND) or cerebral infarction, with median FI 3.5 (IQR 2-8.75) and median FQ 0.015 (IQR 0.01-0.38). Three trials looked at clinical outcome using an outcome scale, with median FI 3 and median FQ 0.011.

Trials reporting on pharmacological interventions had a median FI of 3 (15 trials, IQR 0-5) and median FQ of 0.011 (IQR 0-0.034). Trials reporting on procedural interventions had a median FI of 5 (4 trials, IQR 2-30.5) and median FQ of 0.018 (IQR 0.012-0.088).

### Study quality assessment

Results of study quality assessment are shown in Table 2 (Cochrane risk of bias tool 2.0). Three studies (17.6%) were rated overall as low risk of bias, six studies (35.3%) as some concerns, and eight studies (47%) as high risk of bias.

**Table 2:**
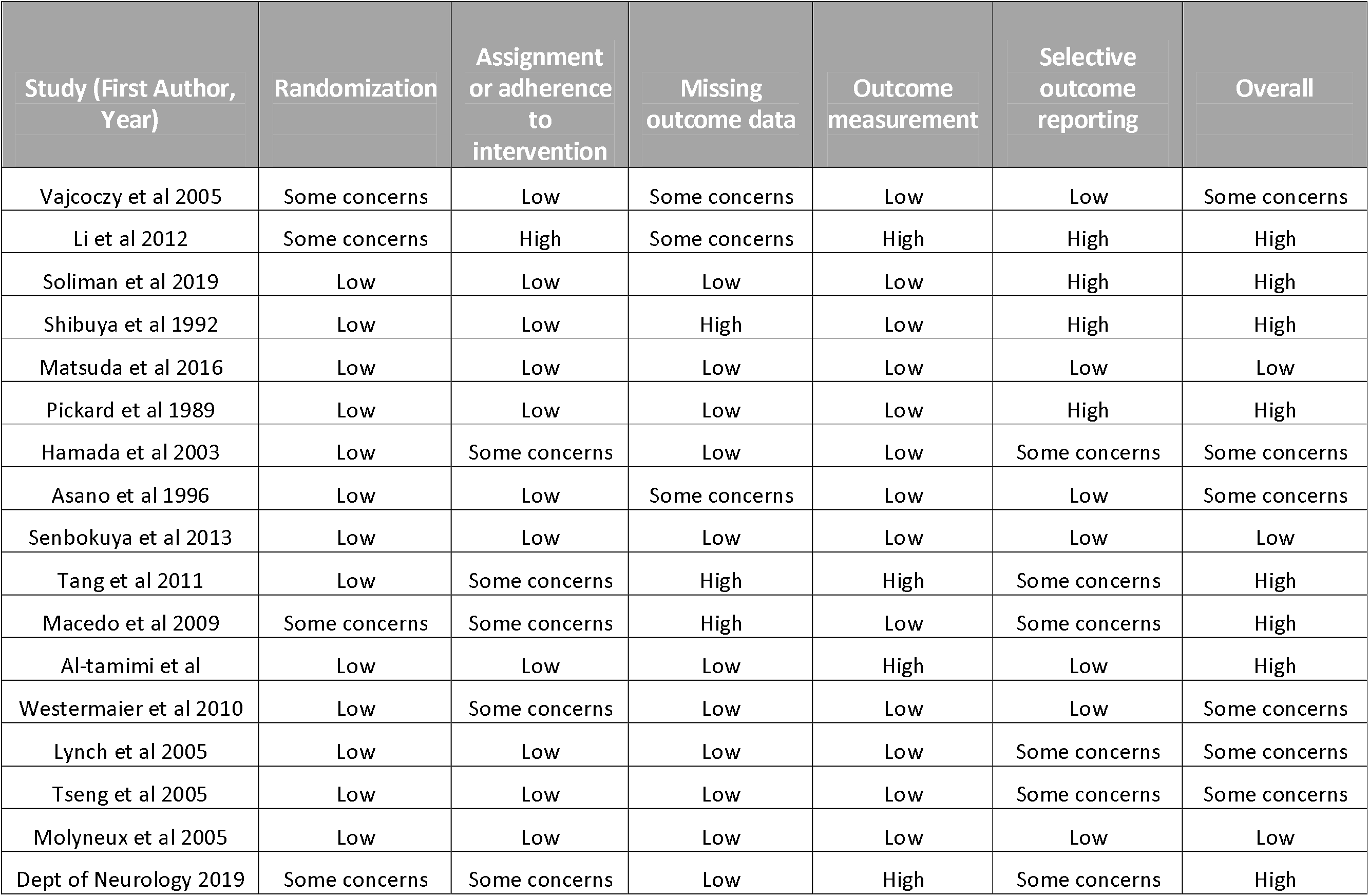
Risk of bias assessment for randomised trials using the Cochrane risk of bias tool 2.0

## Discussion

The median fragility index in our systematic appraisal of randomized trials in aSAH is 3, with an interquartile range of 0 to 5. This is comparable to findings in critical care generally[4] and similar to a previous analysis of cerebrovascular trials[7], but is lower than that in cardiovascular studies, where the median fragility index was 13[5]. The fragility quotient relates the FI to the size of the trial and can be interpreted as the number of patients per 100 required to experience a different event to render the result non-significant. Our analysis shows a median FQ of 0.012, or 1.2 patients per 100, a figure comparable to trials supporting VTE guidelines[28], higher than in cardiovascular trials[5] and not previously reported in trials of aneurysmal subarachnoid haemorrhage.

Six of nineteen trial outcomes (31.6%) are extremely fragile with an FI of 0, meaning simply recalculating the result using Fisher’s Exact Test is sufficient to render the p-value non-significant without any switch in patient events. These findings show that conclusions of trials in aSAH rest on a very low number of events, a fact perhaps not appreciated by clinicians and indicative of a weakness in the trial not apparent from the p-value alone.

Analysis by outcome does not show that fragility is notably different in one particular type of trial or with one type of outcome. Unlike Khan et al.[5] we did not demonstrate higher FI in pharmaceutical trials. We also note that the majority of trials included in our analysis use a surrogate outcome (e.g. vasospasm) and not a patient centred outcome. Again this suggests that the body of evidence currently used to inform management of aSAH is lacking in crucial respects.

In more than one third of trials the fragility index is exceeded by the number of patients lost to follow up, indicating that the outcomes for lost participants could have changed the result to a non-significant one. It further emphasises that an FI of 3 is low and RCTs in aSAH are fragile. Our finding is in agreement with those of Khan et al, and Adeeb et al, in cardiovascular and cerebrovascular RCTs respectively[5,7]. Taken together this suggests the problem is not restricted to a single speciality within medicine but is generic to clinical research.

Only a minority of trials are at low risk of bias assessed by the Cochrane Risk of Bias Tool 2.0, which provides additional support to the suggestion that the quality of evidence for interventions in aSAH is generally low. We have not formally analysed the relationship between risk of bias and fragility index, but note that studies we consider at high risk of bias have a higher FI but lower FQ than other studies (median 3 vs. 2 and 0.011 vs. 0.018 respectively). The lack of a clear relationship indicates that FI analysis provides additional information over and above to usual methods.

Our study has the following strengths: we use a systematic search with no restriction by date or database and analyse the risk of bias alongside fragility to provide a more rounded assessment of trial evidence than p-value alone. We have identified and analysed more trials than previous work in the area. Limitations of our study include exclusion of key trials that do not fit the strict inclusion criteria for fragility analysis, a recognised drawback for the technique. We acknowledge that this drawback means fragility analysis cannot be recommended for all studies at present, although identifying ways to extend the method to studies with multiple groups or non-dichotomous outcomes is an avenue for research.

The number of trials we exclude is a high proportion of all studies identified by our search. While this might be taken as a limitation as above, we believe that as a large number of these exclusions were for methodological weaknesses (e.g. failing to specify a primary outcome in advance) this supports our general conclusion that overall the evidence supporting practice in aSAH is weaker than that suggested on the basis of p-values alone.

We must stress that we do not propose fragility analysis is used in isolation to dismiss trials from consideration, especially as there are no defined thresholds for a low FI or FQ, but that it supplements the usual methods. Lower FI values, particularly where the loss to follow up exceeds the FI, might help flag up concerns about trial reliability and act as an additional gauge of uncertainty of effect. The FQ represents another useful tool to further assess effect strength – consider 2 trials, one with an FI of 6 and an FQ of 0.2 (n=30), and the other with an FI of 30 and an FQ of 0.015 (n = 2000). The higher FI of the second trial might indicate it is more robust, but its lower FQ suggests it may well be less robust than the first in relation to its sample size.

In conclusion fragility analysis is a useful adjunct to critical appraisal of randomised controlled trial evidence in aneurysmal subarachnoid haemorrhage. We caution against over-reliance on p-values and a false dichotomy into “positive” and negative” trials and call for future research to investigate extending fragility analysis to more trial designs and to define thresholds for interpretation.

## Data Availability

Data available on application to corresponding author.

## Supplemental content

**Supplemental Table 1**: Systematic review search strategy designed in Medline

**Supplemental Table 2**: Table of study characteristics

