## Supplemental Table 1 for "The fragility index in randomised controlled trials of interventions for aneurysmal subarachnoid haemorrhage: a systematic review"

**Supplemental Table 1**: Systematic review search strategy designed in Medline

| **#** | **Search term** | **Results** |
| --- | --- | --- |
| 1 | SUBARACHNOID HEMORRHAGE/ AND (exp ANEURYSM/ OR (aneurysm* OR non-trauma* OR spontaneous*).ti,ab) | 11436 |
| 2 | ANEURYSM, RUPTURED/ AND (exp BRAIN/ OR (brain OR cereb* OR intracranial).ti,ab) | 4245 |
| 3 | INTRACRANIAL ANEURYSM/ AND (RUPTURE, SPONTANEOUS/ OR (ruptur* OR burst*).ti,ab) | 8487 |
| 4 | (((subarachnoid OR arachnoid) ADJ6 (haemorrhage* OR hemorrhage* OR bleed* OR blood*)) ADJ6 (aneurysm* OR non-trauma* OR spontaneous*)).ti,ab | 8592 |
| 5 | (((brain OR cereb* OR intracranial) ADJ3 aneurysm*) ADJ3 (ruptur* OR burst*)).ti,ab | 4449 |
| 6 | ((cerebral OR intracranial OR cerebrovascular) ADJ6 (vasospasm OR spasm)).ti,ab | 4903 |
| 7 | (1 OR 2 OR 3 OR 4 OR 5 OR 6) | 22679 |
| 8 | (randomized controlled trial).pt | 499184 |
| 9 | exp "RANDOMIZED CONTROLLED TRIALS AS TOPIC"/ | 133149 |
| 10 | (controlled clinical trial).pt | 327387 |
| 11 | (random*).ti,ab | 1096883 |
| 12 | RANDOM ALLOCATION/ | 101871 |
| 13 | exp "CLINICAL TRIALS AS TOPIC"/ | 336129 |
| 14 | (trial).ti | 213450 |
| 15 | (8 OR 9 OR 10 OR 11 OR 12 OR 13 OR 14) | 1606780 |
| 16 | exp ANIMALS/ NOT HUMANS/ | 4664145 |
| 17 | (review).pt | 2614671 |
| 18 | (meta analysis).pt | 110298 |
| 19 | (news).pt | 753887 |
| 20 | (comment).pt | 829980 |
| 21 | (editorial).pt | 518240 |
| 22 | ("cochrane database of systematic reviews").so | 14683 |
| 23 | ("systematic review" OR "literature review").ti | 150915 |
| 24 | (16 OR 17 OR 18 OR 19 OR 20 OR 21 OR 22 OR 23) | 9028906 |
| 25 | 15 not 24 | 681292 |
| 26 | (7 AND 25) | 517 |
