## Supplemental Table 2 for "The fragility index in randomised controlled trials of interventions for aneurysmal subarachnoid haemorrhage: a systematic review"

**Supplemental Table 2**: Table of study characteristics

| **Study** | **Setting** | **n** | **Intervention** | **Comparator** | **Primary outcome** | **Primary outcome result** | **p value** |
| --- | --- | --- | --- | --- | --- | --- | --- |
| Vajkoczy et al 2005 | Multi-centre, Germany | 32 | Clazosentan | Placebo | Vasospasm | Clazosentan: 6/15  Placebo 15/17 | 0.008 |
| Li et al 2012 | Single centre, China | 63 | Naomai jiejing decoction 1 and 2 | Usual care | Vasospasm Re-bleeding  Hydrocephalus | NJD 9/31; control 17/32 NJD 1/31; control 2/32 Hydrocephalus non significant | <0.05 |
| Soliman et al 2019 | Single centre, Egypt | 90 | Magnesium sulphate | Milrinone | Vasospasm | Mag: 9/45 Mil: 21/45 | 0.007 |
| Shibuya et al 1992 | Multi-centre, Japan | 276 | AT877 | Placebo | Angiographic Vasospasm Symptomatic vasospasm Poor outcome using GOS | AT 36/95; placebo 59/96 AT 43/123; placebo 64/129 AT 12/99; placebo 29/110 | 0.0023  0.0247  0.0152 |
| Matsuda et al 2016 | Multi-centre, Japan | 148 | Cilostazol | Placebo | Symptomatic vasospasm | Cilostazol: 8/74 Placebo: 18/74 | 0.031 |
| Pickard et al 1989 | Multi-centre, UK | 554 | Nimodipine 60mg PO/NG 4 hourly | Placebo | Cerebral infarction | Nimodipine: 61/278 Placebo: 92/276 | 0.014 |
| Hamada et al 2003 | 2 centres, Japan | 110 | Microcatheter intrathecal urokinase infusion into cisterna magna | Usual care | Symptomatic vasospasm | ITUKI: 5/53 Control: 16/57 | 0.012 |
| Asano et al 1996 | Multi-centre, Japan | 162 | ASV (free radical scavenger) | Placebo | DINDs | AVS: 27/80 Placebo: 39/82 | <0.05 |
| Senbokuya et al 2013 | Multi-centre, Japan | 109 | Cilostazol 100mg BD 14 days | Usual care | Symptomatic vasospasm | Cilostazol: 7/54 Control: 22/55 | 0.0021 |
| Tang et al 2011 | Single centre, China | 150 | Hyperbaric oxygen therapy | Usual care | GOS 4-5 | HBO2: 54/60 Control: 46/60 | <0.05 |
| Macedo et al 2009 | Single centre, Brazil | 21 | Simvastatin 80mg nocte | Usual care | Mortality  Vasospasm | SVT 2/11, control 6/10 SVT 1/11, control 4/10 | unknown |
| Al-tamimi et al 2012 | Single centre, UK | 210 | Medtronic lumbar drain | Usual care | DIND | Lumbar drain: 22/105 Control: 37/105 | 0.021 |
| Westermaier et al 2010 | Single centre, Germany | 110 | MgSO4 to achieve serum concentration 2-2.5mmol/l | Normal saline | Delayed ischaemic infarction | MgSO4: 12/54 Control: 27/53 | 0.002 |
| Lynch et al 2005 | Single centre, USA | 39 | Simvastatin 80mg nocte | Placebo | Vasospasm | SVT: 5/19 Placebo: 12/20 | <0.05 |
| Tseng et al 2005 | Single centre, UK | 80 | Pravastatin 40mg | Placebo (lactose) | Vasospasm | Prav: 17/40 Placebo: 25/40 | 0.006 |
| Rao et al 2019 | Single centre, China | 72 | Continuous stellate ganglion block | Usual care | Cerebral Vasospasm | block 5/36; control 22/36 | <0.05 |
| Molyneux et al 2005 | Multi-centre, Europe, North America | 2143 | Endovascular treatment | Surgical treatment | MRS 3-6 at 1 year | coil 250/1063 neurosurg 326/1055 p=0.0001 | 0.0001 |
